# The Context of Recovery Among Adults Who Recover from Myalgic Encephalomyelitis/Chronic Fatigue Syndrome and Post-COVID Condition Following Mind-Body Approaches: A Qualitative Descriptive Study Protocol

**DOI:** 10.64898/2026.09.24.26363855

**Authors:** Ligia Cordovani, Lujain Almazyad, Dena Zeraatkar, Jason W Busse

## Abstract

**Background:** Myalgic encephalomyelitis/chronic fatigue syndrome (ME/CFS) and post-COVID condition are complex, debilitating illnesses characterized by persistent fatigue and post-exertional malaise. Recovery is uncommon without targeted intervention; however, emerging clinical reports suggest that a subset of patients experience full resolution of symptoms after engaging in mind-body approaches. Patient perspectives on these interventions are divided, and qualitative evidence exploring successful recovery pathways is rare. This protocol outlines a study to investigate the contextual factors and patient-constructed explanations surrounding full recovery from ME/CFS or post-COVID condition using mind-body approaches.

**Methods and analysis:** We will employ a qualitative descriptive design rooted in a constructivist paradigm. Purposive and snowball sampling will be used to recruit approximately 20 adults from established infection-associated fatigue networks who self-report full recovery (lasting three months or more) from ME/CFS or post-COVID condition following a mind-body approach. Data will be collected through semi-structured interviews conducted via Zoom, and investigator reflexivity will be reported. Demographic information, including comorbidities, will be documented to contextualize the sample. Data will be analyzed inductively using reflexive thematic analysis. To ensure research rigor and reporting transparency, the study will adhere to the 15-point checklist of criteria for good thematic analysis proposed by Braun and Clarke.

**Dissemination:** We will publish our findings in a peer-reviewed, open-access journal and present at relevant academic conferences. A lay summary of the results will also be shared directly with participating patient recovery networks.

## Introduction

Myalgic encephalomyelitis/chronic fatigue syndrome (ME/CFS) and post-COVID condition (also known as long COVID) are complex, debilitating illnesses that frequently follow an infectious trigger and share overlapping symptom profiles, including persistent fatigue, post-exertional malaise, cognitive dysfunction, and autonomic instability [1–4]. The point prevalence of post-COVID condition between 2023 and 2024 has been estimated between 2.9% and 6.7% [5, 6], whereas ME/CFS is estimated to affect 0.4–1% of the general population [7, 8]. Women are affected with either condition 2-3 times more often than men [1, 2, 7, 8]. The etiology of these disorders remains poorly understood, though proposed mechanisms include immune dysregulation, autonomic nervous system dysfunction, and maladaptive brain-body signaling pathways [8–10]. Diagnosis is based on patient-report, and neither condition has established biomarkers [11–14].

Some ME/CFS patients report gradual improvement over time, but without targeted intervention full recovery is achieved in approximately 5% of cases [15]. Full recovery from post-COVID condition in general has been estimated at 7.6% [16]. However, a third of individuals report >75% recovery with specialized care [18]. Emerging clinical reports of recovery have surfaced from individuals who engaged in mind-body approaches such as Cognitive Behavioral Therapy (CBT), Pain Reprocessing Therapy (PRT), and the Lightning Process (LP) [17–23]. These approaches aim to modulate central processes hypothesized to sustain chronic symptoms, including heightened threat perception, symptom hypervigilance, and dysregulation of the autonomic nervous system [10].

A living systematic review of interventions for long COVID found moderate-certainty evidence that CBT likely reduces fatigue and improves concentration and that combined physical and mental health rehabilitation likely improves overall health and reduces depressive symptoms [24]. Additionally, a meta-analysis of individual patient data from eight trials that enrolled 1,298 participants with ME/CFS reported reductions in fatigue, functional impairment, and physical limitations after CBT-based interventions [25]. However, some patients report resistance to mind-body approaches, in part because of concerns that such interventions trivialize or psychologize their symptoms [26].

Studies exploring the experiences of individuals who report having recovered are rare, especially in terms of contextual factors that led to their willingness to engage with mind-body approaches and how participants construct explanations for their recovery following successful treatment [22]. Understanding the perspectives and experiences of those who report recovery from mind-body interventions could inform strategies to support others living with ME/CFS or post-COVID condition.

### Purpose Statement and Research Question

This study will explore the context of recovery among adults who report full recovery from ME/CFS or post-COVID condition following mind-body approaches. Specifically, we will examine: (1) participants’ initial perceptions of these treatments (including resistance), (2) how they came to engage with them, (3) what expectations were set for patients when they engaged in their mind-body approaches (4) how participants construct explanations for perceived recovery following engagement with mind-body approaches, (5) their experiences sharing recovery within patient communities, (6) what recovery means to them, and (7) their perspectives on what might support others still living with ME/CFS or post-COVID condition to consider engaging with mind-body approaches.

Our overarching research question is: What can we learn about the context of recovery among adults who report full recovery from ME/CFS and post-COVID condition following mind-body approaches?

By capturing these experiences, our study aims to contribute to a deeper understanding of recovery pathways and inform future strategies to support patients with ME/CFS and post-COVID condition.

## Methodology

### Study Design

We will employ a qualitative description approach, a methodology well suited to health care research that seeks to provide direct descriptions of participants’ experiences and perceptions to acquire new insights founded in existing knowledge [27, 28]. Qualitative description is particularly appropriate when the goal is to stay close to the data and to the surface of participants’ words and events, yet producing findings that are detailed and nuanced interpretive, accessible, and meaningful [29]. In this study, realities will be presented through people who report recovery from ME/CFS and post-COVID condition following mind-body approaches.

### Study timeline

Data collection has not yet started. Our proposed study timeline is outlined below:

Months 1–2: Pilot test and finalize the interview guide; prepare recruitment materials.

Months 3–8: Recruit participants, conduct interviews, and undertake concurrent data analysis.

Months 9–11: Complete data analysis, refine themes, and synthesize findings.

Months 12–14: Prepare and submit the study manuscript for publication.

This timeline allows for iterative data collection and analysis, consistent with qualitative research methodology, while ensuring adequate time for rigorous interpretation and dissemination of findings.

### Theoretical framework

Our theoretical foundation is informed by a growing body of research demonstrating the potential benefits of mind-body approaches for managing chronic symptoms associated with ME/CFS and post-COVID condition [20, 21]. These approaches, which emphasize the interaction of physiological, psychological, and behavioral processes, have shown potential in supporting symptom self-management, improving quality of life, and enhancing coping strategies among individuals living with persistent and often debilitating symptoms. In addition to empirical evidence, our framework is informed by reports from patient communities, including individuals living with ME/CFS, where individuals have described benefits from engaging in mind-body practices, including improvements in symptom management, emotional well-being, and daily functioning [22].

### Philosophical foundations

We will adopt a constructivist paradigm, which assumes that individuals create meaning through their interactions with the world and that multiple realities exist as a result of these experiences. Ontologically, our study will be grounded in relativism, recognizing that reality is subjective and shaped by individual perspectives. Epistemologically, we align with subjectivism, acknowledging that understanding is co-created through participants’ interpretations of their experiences and the researcher’s involvement in the inquiry process. Accordingly, knowledge will be co-constructed with participants through semi-structured interviews guided by flexible, open-ended questions that allow the interviewer to explore emerging perspectives and novel insights. Consistent with a constructivist approach, data will be analyzed inductively to identify patterns and meanings arising from participants’ accounts. To support a constructivist axiology, we (investigators) will engage in reflexive practices by documenting and critically examining our assumptions, values, and interpretations throughout the study. Finally, our constructivist rhetoric will be reflected in a narrative style of reporting that prioritizes participants’ voices and employs language and terminology appropriate to qualitative health research [29, 30]

### Setting and participants

Our study will be conducted at the Michael G. DeGroote National Pain Centre at McMaster University, Hamilton, Ontario, Canada. Eligible participants will be: adults (>18 years old) who have fully recovered for ≥3 months (according to a self-report) from ME/CFS or post-COVID condition through a mind-body approach (e.g., CBT, PRT, mindfulness, graduated activity), are English-speaking, and are able to provide informed consent.

### Recruitment

Participants will be recruited via networks of people who report recovery from ME/CFS or post-COVID condition (e.g., Recovery Norge [https://www.recoverynorway.org/], the Oslo Chronic Fatigue Network [https://www.oslonetwork.no/], the International Collaborative on Fatigue Following Infection [https://www.coffi-collaborative.com/]). These networks will be contacted and asked to circulate notice about our study to their membership through channels they deem appropriate (e.g., newsletters, websites, social media profiles). Advertisements will be circulated electronically. Interested individuals will contact a research assistant by email. Participants identified through snowball sampling will be provided with the research assistant’s contact information by their referrers and will initiate contact if interested. Participants will be offered a gift card as compensation for their time.

### Sampling

We will use a non-probabilistic sampling, as the intent of qualitative research is not to generalize results [31]. We will employ purposive sampling to recruit participants with direct experience of full recovery from ME/CFS or post-COVID condition using a mind-body approach [30, 31]. Snowball sampling will supplement recruitment by encouraging participants to circulate advertisements within their networks [31]. We will use maximum variation sampling to seek diversity of viewpoints; for example, approximately equal numbers of participants will be sought for ME/CFS and post-COVID condition [28].

Our sample size will be informed by the concept of information power, which holds that the more information the sample provides relevant to the study’s research question, the fewer participants are needed [32]. Given the study’s focused aim, specific population, established theoretical framework, and use of in-depth interviews, a relatively small sample per condition (approximately 10 participants) may be sufficient. Therefore, we estimate that a total sample of approximately 20 participants will be required to attain meaningful description. However, the adequacy of the final sample size will be evaluated continuously during the research process by the research team, findings will be regularly discussed, and the information power will be repeatedly appraised and supported by preliminary analysis and emergent analytic ideas [33].

### Data Collection

We will collect data through semi-structured individual interviews conducted through the Zoom videoconferencing platform. (See Appendix I for our Interview guide) The interview guide will be piloted for feasibility and acceptability with two participants, who will not be part of the study sample. Their input might inform minor modifications to question order and clarity. Study participants will be invited to describe their perceptions of factors that contributed to their recovery. All interested participants will receive an electronic informed consent form for review, operationalized through REDCap, prior to the interview. The co-investigator (LC), who is an anesthesiologist with extensive experience in qualitative data collection, will conduct all interviews. Interviews will be approximately 45 minutes in duration.

Demographic data (age and presence of comorbidities such as fibromyalgia, depression or anxiety, irritable bowel syndrome, sleep disorders, and orthostatic intolerance) will be collected during interviews, as these factors have been described as possible predictors of recovery in patients with ME/CFS and post-COVID condition [34–36]. The purpose of collecting demographic information is to characterize the study sample and contextualize the scope and transferability of the findings. Consistent with qualitative research methodology, demographic characteristics will not be analyzed as variables or used for comparison or interpretation.

Recording will include audio only; participants will be offered the option to turn their cameras off at any point during the interview. Data collection and analysis will occur concurrently, and the investigators (LC, DZ, JWB, and LA) will meet weekly to discuss interview findings and make minor modifications to the interview guide, if needed.

### Data Analysis

To analyse the data, we will use a reflexive thematic analysis approach, a method that prioritizes the values of qualitative paradigms (i.e., a non-positivist framework) and foregrounds subjectivity and interpretation [37, 38]. This approach includes six steps: (1) transcribing the interviews and listening to the voices of the participants repeatedly, (2) a structured coding process, (3) sorting out codes into initial themes, (4) reviewing potential themes, (5) defining and naming themes, and (6) producing a report addressing the research question and going beyond a simple description [37]. (Figure 1)

**Figure 1.**
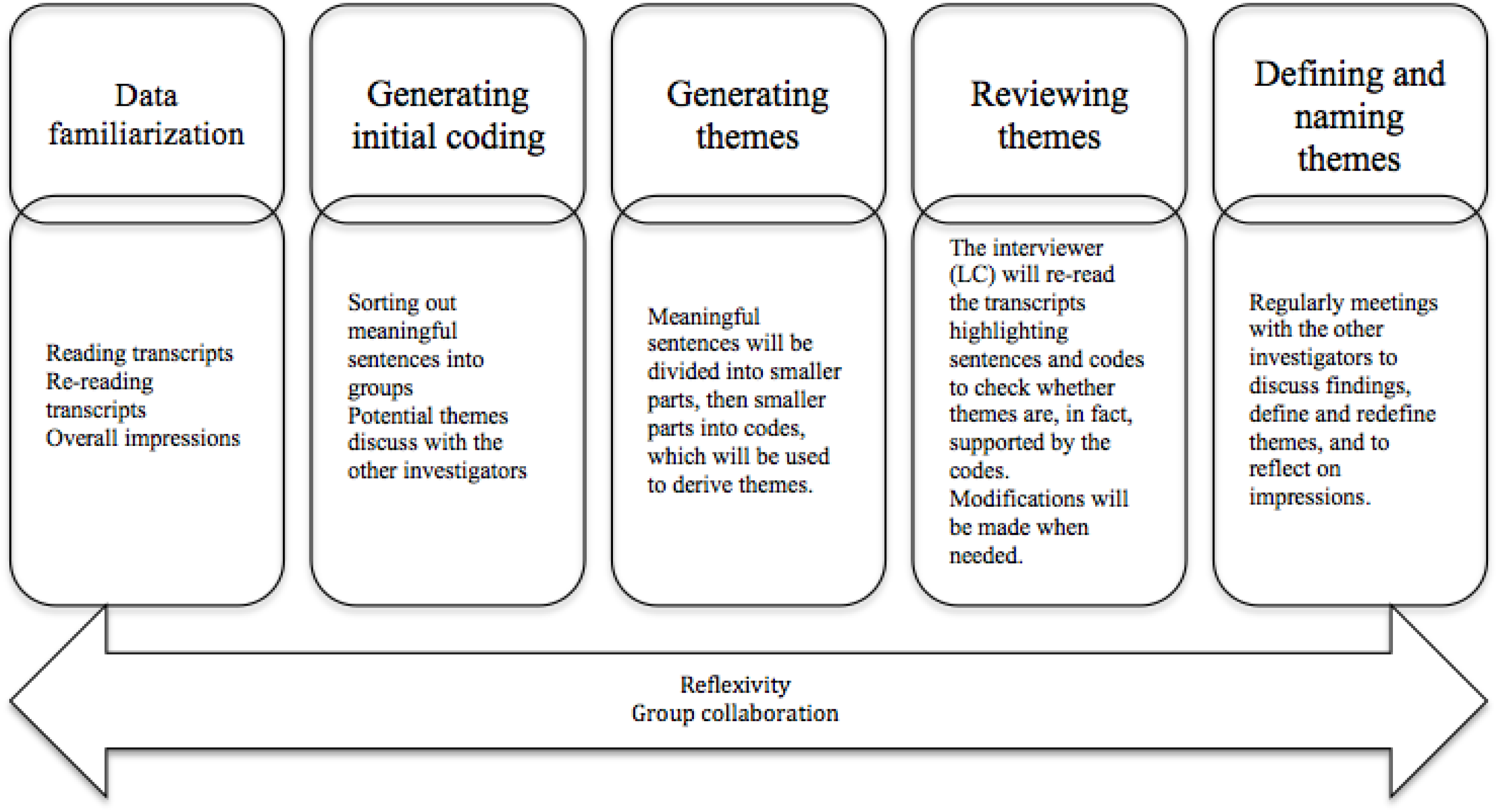
Reflexive thematic data analysis process. Adapted from Braun and Clarke and Campbell et al.

Themes, as the final analytic product, will be organized in tables to create a visual and contextual interpretation. We will follow the core assumptions of reflexive thematic analysis as outlined by Braun and Clarke [37]: (1) researcher subjectivity is considered a resource for knowledge generation; (2) analysis and interpretation should be insightful, thoughtful, and rich; (3) data analysis involves both immersion for engagement and distancing for reflection; (4) a single coder with group collaboration (rather than consensus coding) is recommended; (5) reflexivity is essential to high-quality analysis; and (6) data analysis is a creative process within a framework of rigor grounded in methodological, theoretical, and philosophical underpinnings, meaning multiple analyses are possible and the researchers must determine which best serves the project [37, 39].

We will document coding decisions, theme development, and analytic reflections throughout the analysis using analytic memos to provide a transparent record of the evolving interpretation of the data. Emerging interpretations and themes will be discussed during regular meetings with the study investigators to enrich interpretation rather than achieve coding consensus.

### Strategies to Enhance Rigor

We will follow the 15-point checklist of criteria for good thematic analysis proposed by Braun and Clarke [37, 39]. The data will be transcribed in detail and checked against the original recordings to ensure accuracy. During analysis, each data item will receive equal attention, and all relevant data extracts will be systematically coded. Themes will be developed to be coherent, consistent, and clearly distinctive from one another. The resulting analysis will present a well-organized and compelling account of the data in relation to the context of recovery from ME/CFS and post-COVID condition, demonstrating a clear analytic narrative rather than a collection of disconnected themes. The thematic analysis approach will be transparently described and justified, with explicit attention to the analytic procedures undertaken. Throughout the process, the study team (LC, DZ, JWB, LA) will be positioned as active agents in knowledge construction, acknowledging our interpretive role in generating and refining the themes.

Additionally, we will pay attention to criteria to enhance study rigor, specifically those proposed for qualitative descriptive approaches [27]. Credibility will be enhanced through our deep engagement with the data, active involvement throughout the research process, methodological rigor, and ongoing collaboration among the research team. Transferability will be supported by providing rich descriptions of the study context, participants, and data, as well as illustrative participant quotations. We will strengthen dependability and confirmability through data documentation, transparent reporting of the study methods, and peer debriefing to examine and refine interpretations and findings. Furthermore, one of the co-investigators (LC) will keep a reflective journal throughout the study. Alongside regular investigator meetings, this journal will support ongoing reflexivity by helping the research team identify, reflect on, and critically examine how our positions, experiences, beliefs, and perceptions of the phenomenon under study influence both the research process and the resulting findings.

### Data Availability

Following completion of the study, qualitative data will be managed in accordance with participant consent and institutional ethics requirements. Qualitative interview data contain sensitive personal narratives that cannot be fully de-identified without compromising participant confidentiality [41]. Therefore, raw transcripts will not be made publicly available. De-identified thematic summaries and illustrative quotations will be included in the published manuscript.

Requests for access to additional de-identified data may be submitted to the first author (LC), where they will be reviewed in accordance with institutional ethical requirements.

### Patient and Public Involvement

The interview guide will be piloted with two members of the public (one with ME/CFS condition and one with post-COVID condition). Their input will be used to inform modifications, if needed. Additionally, our study is informed by prior qualitative work that centered on patient perspectives on recovery from ME/CFS [21], and our study team members include experts in chronic pain research and management.

### Ethical and safety considerations

Our study has received ethics approval from the Hamilton Integrated Research Ethics Board (HiREB) at McMaster University (#19531) and will be conducted in accordance with the Tri-Council Policy Statement: Ethical Conduct for Research Involving Humans (TCPS 2). Verbal consent will be obtained from all participants. Participants will receive a copy of an electronic informed consent form for review, operationalized through REDCap, before their interview. At the beginning of the interview, the interviewer will discuss the study, confirm the participant has read the informed consent form, and provide an opportunity to ask questions. After this is complete, verbal consent will be acquired and the study investigator will document this on the consent form.

To protect participants’ privacy, all responses will remain confidential and transcripts will not be linked to participants’ names, only to participant ID numbers. Transcripts will be downloaded from the Zoom platform after each interview, de-identified by the interviewer (LC), and stored in a password-protected file. All study data will be saved remotely in accordance with our University data storage policy. Co-investigators (LC, LA, DZ and JWB) will have access to de-identified transcript fragments to facilitate discussion of findings. Recordings will be permanently deleted after transcripts are downloaded. All included quotes will be carefully de-identified in our manuscript and reviewed to reduce risk of deductive identification. Moreover, participants may withdraw from the study at any time prior to or during the interview. Data withdrawal will be permitted for a period of one month following interview completion, after which data will be incorporated into analysis and cannot be withdrawn.

Additional measures will be taken to minimize participants’ risk and discomfort during data collection. Interviews will be conducted via the Zoom videoconferencing platform; however, only audio recordings, and not video recordings, will be collected. To support participant comfort and autonomy, individuals may choose whether to keep their camera on or off during the interview. Zoom is an externally hosted, cloud-based service that has been approved by our Research Ethics Board for use in this study. As with any data collected through external servers, there is a small risk of a privacy breach. This risk is outlined in the participant information letter. Participants who prefer not to use Zoom may choose an alternative interview format, such as a telephone interview.

### Dissemination plan

We will submit our study findings for publication in a peer-reviewed open-access journal and present at relevant academic conferences. A lay summary of findings will be shared with participating networks and communities.

### Strengths and Limitations

Our study has several anticipated strengths. First, it addresses a significant gap in the literature by exploring the lived experiences of individuals who report full recovery from ME/CFS or post-COVID condition through mind-body approaches; a topic that has received limited investigation. Second, the use of qualitative description provides a methodologically appropriate framework for generating clinically accessible findings that stay close to participants’ own words and experiences [27, 28]. Third, the use of reflexive thematic analysis, with the rigorous framework of Braun and Clarke, gives analytical depth with transparency and reflexivity [37–39]. Fourth, the recruitment strategy uses existing recovery networks and maximum variation sampling to gather a range of perspectives from both people who have recovered from ME/CFS and post-COVID condition.

There are certain limitations that must be acknowledged. Our study is predicated on self-reported recovery, which may have a potentially heterogeneous meaning, as there is no consensus definition of "full recovery" from ME/CFS or post-COVID[42]. This heterogeneity will be investigated as part of our analysis. The self-selection of participants into the study may represent a subset of individuals who have positive perceptions of treatment and successful outcomes as a result of recruitment through recovery-focused networks (e.g. Recovery Norge, COFFI) and snowball sampling. Such networks may mirror ideological clustering around mind-body methods and exclude those who tried but did not recover or who recovered via other means. Additionally, qualitative studies typically involve smaller sample sizes than quantitative studies, which may result in a narrower range of perspectives and experiences being captured. Consequently, the findings will not be statistically generalizable. However, they will provide transferable insights that can inform future research and clinical practice. Transferability will be enhanced through the inclusion of rich, detailed descriptions of participants’ contexts and experiences, facilitating readers to determine the applicability of the findings to their own settings and circumstances.

## Authors’ contributions

Jason W. Busse (Principal Investigator): conceptualization and review.

Ligia Cordovani (Co-Investigator): conceptualization, methodology, writing, review, and editing.

Lujain Almazyad (Co-Investigator): review and editing.

Dena Zeraatkar (Co-Investigator): conceptualization and review.

## Ethics Statement

This study has received ethics approval from the Hamilton Integrated Research Ethics Board (HiREB) at McMaster University (#19531).

## Data Availability

No datasets were generated or analysed during the current study. All relevant data from this study will be made available upon study completion.

